# Implementation of SMS and voice message reminders to reduce childhood immunization dropout rate in urban settings: A Pilot Study in Lomé-Togo in 2026

**DOI:** 10.64898/2026.04.19.26350799

**Authors:** Soliou Badarou, Kokou Mawunya Attah, Kokou Herbert Gounon, Aimé Serge Dali, Xavier Richard Sire, Erinna Corinne Dia

**Affiliations:** Unicef, United Nations Children’s Fund, Togo Country Office; Department of Public Health, University of Lomé, Faculty of Health Sciences, Lomé, Togo; African Center for Research in Epidemiology and Public Health, Lomé, Togo; Division of Health System and Policy, Institut National de Santé Publique, Abidjan, Côte d’Ivoire; Department of Global Health, University of Washington, Seattle, WA, USA

**Keywords:** SMS reminder, Digital, mHealth, Dropout rate, Feasibility, Expanded Program on Immunization, Togo

## Abstract

**Objective:** This study aimed to assess the effectiveness of SMS and voice message reminders in reducing the dropout rate in Lomé-Togo, in 2026.

**Methods:** We conducted a cross-sectional study between October 2025 and March 2026 in the Grand Lomé region. The intervention consisted of an integrated digital system used by health facilities to send automated SMS. Categorical variables were described in terms of frequency and proportion; Fisher’s exact test was used to compare proportions. Quantitative variables were described by their means accompanied by their standard deviation; the Wilcoxon rank-sum test was used to compare means. The significance level for statistical tests was set at 5%.

**Results:** A total of 30 health facilities were included. Seventy percent (70.0%) of the health facilities used messages associated with calls. Ninety percent (90.0%) of participants found the reminders useful, and 60.0% reported an improvement in Expanded Program on Immunization services related to their use. Among participants who received a reminder, 51.0% kept their vaccination appointments. The Penta 1/3 dropout rate decreased from 3.2% before the intervention to 1.3% (p < 0.001). Among the 323 parents of children included, only 20.74% reported receiving a reminder by phone. Sixty-point-five percent (60.5%) preferred to receive both text messages and voice calls.

**Conclusion:** This study demonstrates the operational feasibility of an SMS/call-based reminder system in reducing dropout rate for childhood vaccination in Togo.

## 1. Introduction

Vaccination is one of the most effective public health interventions. Routine childhood vaccination has prevented an estimated 154 million deaths worldwide between 1974 and 2024, including 146 million among children under five [1]. In 2023, only 84% of children worldwide received the three doses of the diphtheria, tetanus, and pertussis (DTP3) vaccine, leaving an additional 2.7 million children without full protection compared to pre-pandemic levels in 2019 [2]. According to the Global Burden of Disease 2023 analysis, the number of “zero-dose” children reached 15.7 million in 2023, with more than half concentrated in just eight countries (Nigeria, India, the Democratic Republic of the Congo, Ethiopia, Somalia, Sudan, Indonesia, and Brazil), reflecting persistent inequalities [3].

In sub-Saharan Africa, only 54.1% (95% CI: 53.7–54.5%) of children aged 12 to 23 months were fully vaccinated [4]. Furthermore, the prevalence of “zero-dose” children was 12.19% (95% CI: 11.82–12.56) [5].

In Togo, vaccination coverage has increased, raising from 92% for Penta 1 in 2022 to 95.2% in 2024, and from 88% to 90.3% for Penta 3 over the same period; with a dropout rate for Penta1/Penta3 reaching 5% and 30.3% for RR1/RR2 in 2024 [6,7]. These trends are confirmed by field data; a household survey in the Haho district revealed that 15.8% of children aged 12 to 23 months had had no contact with the immunization program [8].

Mobile technologies, which are widely available even in resource-limited settings, offer a promising strategy for reducing vaccine dropout rates. A meta-analysis conducted in Africa showed that mHealth interventions more than doubled the likelihood of children receiving a full course of vaccination (OR 2.15; 95% CI: 1.70–2.72; p < 0.001) [9]. Studies conducted in Nigeria have also demonstrated that telephone reminders offer the advantage of personalized follow-up, with the ability to recontact in case of non-response and answer parents’ questions [10]. However, few studies have evaluated these interventions in the specific context of French-speaking West Africa, and none have been conducted in Togo. This study aimed to assess the effectiveness of SMS and voice message reminders in reducing the dropout rate in Lomé-Togo, in 2026.

## 2. Methods

### 2.1. Study Design, Period, and Setting

We conducted a cross-sectional study in health facilities between October 2025 and March 2026. The study took place in Lomé, Togo, within the Grand Lomé health region.

### 2.2. Inclusion Criteria

We included in the study, on the one hand, all health facilities in the Grand Lomé health region that have a catchment area, a maternity ward, and a vaccination service. On the other hand, we included parents of children born in one of the catchment areas of the selected health facilities starting in October 2025 and who had not yet received their BCG dose.

### 2.2. Method and Sample Size

We conducted an exhaustive recruitment of eligible public health facilities, within which we performed simple random sampling of at least 10 parents of eligible children per facility; this corresponds to a sample size of at least 300 parents.

### 2.3. Data Collection and Analysis

Data collection was performed by reviewing records from DHIS2 (*District Health Information System 2*) [11], the database of the implemented system and using a face-to-face administered questionnaire. The data were analyzed using R software version 4.5.0 [12]. Categorical variables were described in terms of frequency and proportion; Fisher’s exact test was used to compare proportions. Quantitative variables were described by their means accompanied by their standard deviation (SD); the Wilcoxon rank-sum test was used to compare means. The significance level for statistical tests was set at 5%.

### 2.4. The Intervention

The intervention consisted of the development of the “Vaxy-Togo” application, which was implemented and became operational in eligible health facilities starting in October 2025. The solution consists of two main components: a dashboard and a mobile application (**Table 1**). Parents received a confirmation SMS upon enrollment. Parents able to read received an SMS; parents who could not read received a phone call (based on the language they have chosen), and if the call went unanswered, an SMS was sent 24 to 48 hours before the scheduled vaccination date.

**Table 1.** Description of the Intervention.

| Component | Description | Primary Users | Key Features |
| --- | --- | --- | --- |
| **Dashboard** | Online interface for managing, monitoring, and supervising the reminder system | District managers and health facilities coordinators | Overall monitoring of vaccination activities.<br>Supervision of health workers<br>Viewing of aggregated statistics (reminders sent, attendance rates, vaccination coverage) |
| **Mobile App** | Apps installed on a smartphone, used for data entry and tracking in the field | Health workers, midwives, and vaccinators | Enrollment<br>Birth registration<br>Confirmation of children’s attendance at vaccination sessions<br>Generation and tracking of automatic reminders (SMS and voice messages) |

**Table 2.** Characteristics of reminders, Lomé, Togo, 2026 (N = 30)

| Characteristic | Type of reminders |  | Overall | p-value<sup>2</sup> |
| --- | --- | --- | --- | --- |
|  | SMS or call<br>N = 9<sup>1</sup> | SMS and call<br>N = 21<sup>1</sup> | N = 30<sup>1</sup> |  |
| **Frequency of reminders** |  |  |  | 0.111 |
| 24 hours in advance | 4 (44.4) | 3 (14.3) | 7 (23.3) |  |
| 48 hours in advance | 2 (22.2) | 13 (61.9) | 15 (50.0) |  |
| Other | 3 (33.3) | 5 (23.8) | 8 (26.7) |  |
| **Usefulness of reminders** |  |  |  | >0.999 |
| No | 1 (11.1) | 2 (9.5) | 3 (10.0) |  |
| Yes | 8 (88.9) | 19 (90.5) | 27 (90.0) |  |
| **Noticed improvement** |  |  |  | >0.999 |
| No | 4 (44.4) | 8 (38.1) | 12 (40.0) |  |
| Yes | 5 (55.6) | 13 (61.9) | 18 (60.0) |  |

<sup>1</sup> n (%)<sup>2</sup> Wilcoxon rank sum test; Fisher's exact test

### 2.5. Outcomes

The primary outcome is the specific dropout rate (Penta1/3). Secondary outcomes include Penta 3 vaccination coverage, the contact rate, the reminder delivery rate, the appointment adherence rate and parental perceptions. The contact rate is defined as the number of messages/calls sent divided by the number of enrolled phone numbers; the delivery rate as the total number of messages/calls delivered divided by the total number sent; and the appointment adherence rate as the total number of children who have received any type of vaccine divided by the total number who received a reminder.

### 2.6. Ethical and Regulatory Considerations

This study received ethical approval from the Institutional Review Board of the School of Health Sciences at the University of Lomé in Togo [N° 409/2026/CE-FSS/19/01]. All participants signed a consent form. The confidentiality of the data collected was respected. To ensure anonymity, no information that could identify the participants was used.

## 3. Results

### 3.1. Characteristics of Reminders

We included a total of 30 health facilities. Seventy percent (70.0%) of the health facilities used messages associated with calls. Half of reminders (50.0%) were sent 48 hours before the appointment. Ninety percent (90.0%) of health facilities found the reminders useful, and 60.0% reported a perceived improvement of EPI services related to their use (**Tableau 2**).

### 3.2. Penta 1/3 dropout rate and Penta 3 vaccination coverage

The Penta 1/3 dropout rate decreased following the implementation of the intervention. The rate fell from 3.2% before the intervention to 1.3% after the intervention, with a statistically significant difference (p < 0.001) (**Figure 1.A**). Vaccination coverage for Penta 3 increased following the implementation of the intervention. The coverage rose from 105% before the intervention to 108% after the intervention. (**Figure 1.B**).

**Figure 1.**
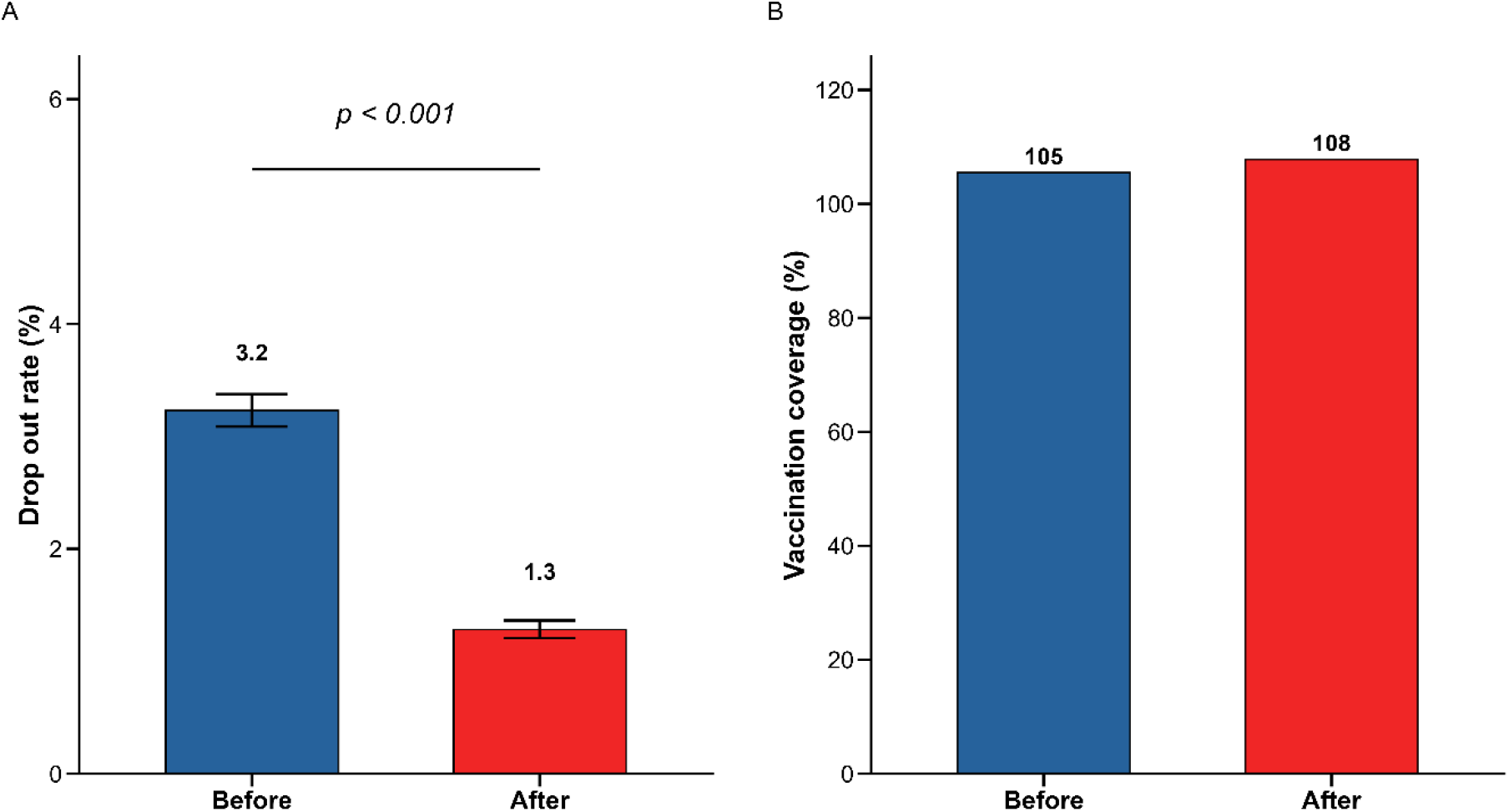
**A**. Penta 1/3 dropout rate (Figure 1.A) and Penta 3 coverage (Figure 1.B) before and after the intervention, Lomé-Togo, 2025

### 3.3. Contact rates, reminder delivery rates, and appointment adherence rates

The contact rate for reminders was 45.0%, while the delivery rate was 47.0%. Among participants who received a reminder, 51.0% kept their vaccination appointments (**Figure 2**).

**Figure 2.**
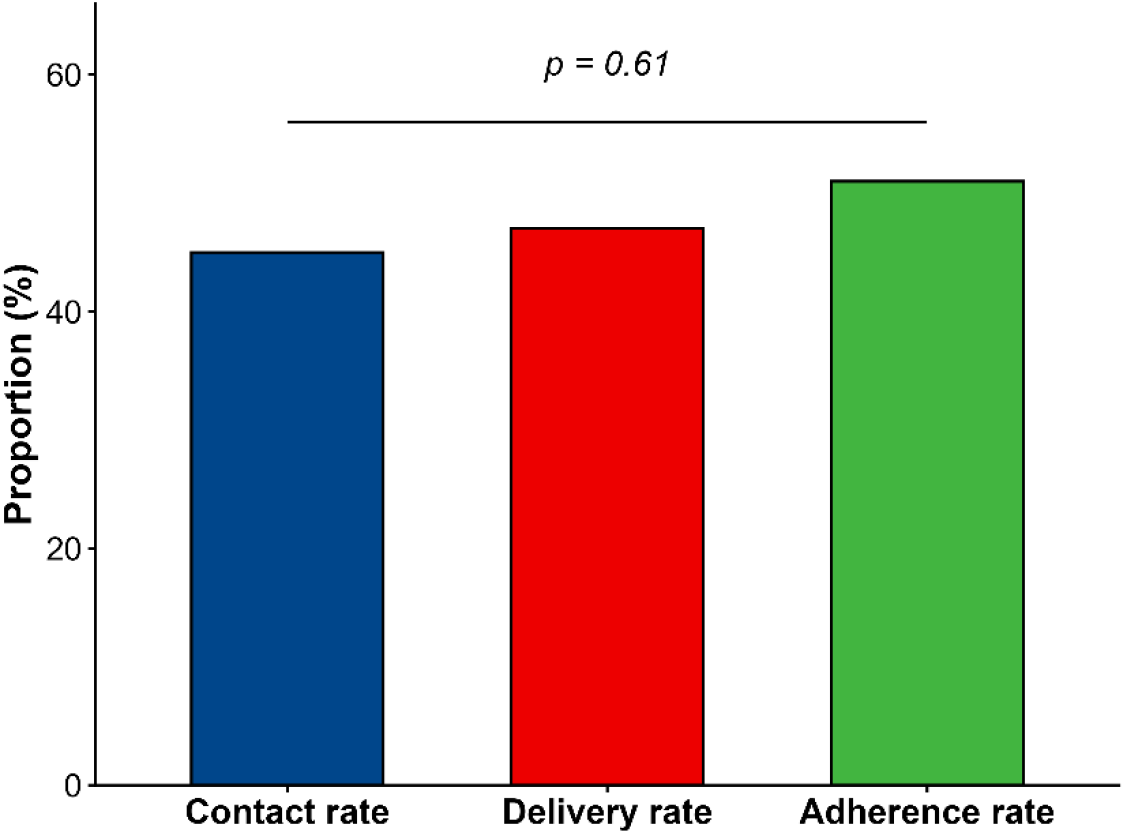
Contact rates, delivery rates, and appointment adherence rates, Lomé-Togo, 2025

### 3.4. Parents’ perceptions of the reminder system

In total, we included 323 parents of children. Among these participants, only 20.74% (N = 67) reported receiving a reminder by phone. Sixty-point-five percent (60.5%) preferred to receive both text messages and voice calls, followed by those who preferred voice calls only (32.4%). The preferred frequency of reminders for every appointment was mainly once (43.7%) or twice (42.3%) (**Table 3**).

**Table 3.**
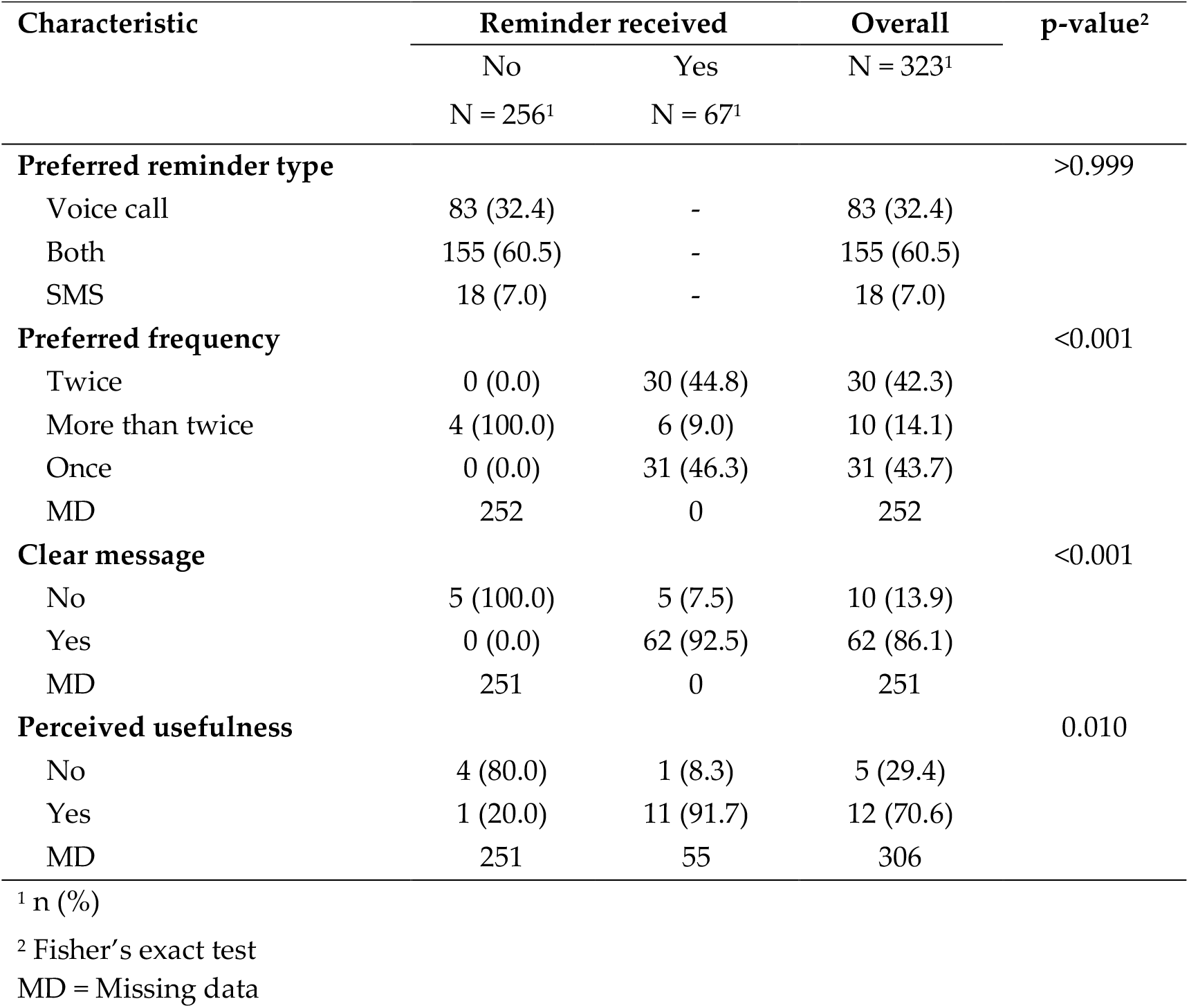
Parents’ perceptions of the reminder mechanism, Lomé, Togo, 2026.

## 4. Discussion

We reported a significant reduction in the Penta 1/3 dropout rate, which fell from 3.2% before the intervention to 1.3% after implementation (p < 0.001). Penta 3 vaccination coverage increased from 105% to 108% over the same period.

### 4.1. Dropout Rate

We reported a Penta 1/3 dropout rate of 3.2% before the intervention and 1.3% after, both of which remain below the 10% threshold that the WHO considers indicative of poor service utilization. Similar results have been reported in a review of published and unpublished literature on SMS-based vaccination reminders in Africa which found that all identified studies reported improvements in either vaccination coverage or dropout rates [13]. In Nigeria, a four-arm randomized trial in Ilorin found immunization completion rates of 99.2%, 99.3%, and 97.0% for the call reminder, SMS reminder, and SMS immunization facts groups, respectively, versus 90.4% for controls [10]. Our similar results suggest that even in a context where baseline dropout rates are already below the WHO threshold, automated reminders can achieve measurable additional gains.

However higher figures have been reported in a systematic review and meta-analysis of studies across sub-Saharan Africa estimated a pooled prevalence of vaccination drop-out at 26.06% (95% CI: 11.59–30.53) [14]. At the regional level, a review of vaccination data across 13 West African countries reported that the DPT1-to-DPT3 dropout rate was high, with a mean of 16.3% across countries [15]. The comparatively low dropout rate in our study can be explained by our urban setting, where access to health facilities is generally better than in rural or remote areas.

### 4.2. Vaccination Coverage

Penta 3 vaccination coverage increased from 105% to 108% in our study. Both values exceed 100%, which is a known limitation of administrative vaccination coverage data [16]. Our coverage figures above 100% can be explained by the possible cross-boundary attendance of populations from outside registered catchment areas, a common phenomenon in urban African settings.

Similar directional increase in coverage has been reported in a systematic review and meta-analysis of SMS reminder interventions in low- and middle-income countries which described a pooled relative risk of 1.16 (95% CI: 1.10–1.21) for improved childhood immunization coverage [17]. In Zimbabwe, a randomized controlled trial reported an immunization coverage at 14 weeks for Penta 3 reaching 95% in the SMS reminder group compared with 75% in the non-intervention group (p < 0.001) [18]. Our modest absolute change observed can be explained by the already-high baseline coverage that limits the room for further measurable improvement.

### 4.3. Reminder Reach and Appointment Adherence

One of the most striking findings of this study is the contrast between the appointment adherence rate observed among parents who received a reminder (51.0%) and the proportion of enrolled parents who actually reported receiving one (20.74%). This gap carries a key message: the primary limitation of the Vaxy-Togo system, at this stage, is not its effectiveness among those it reaches, but its reach itself.

### 4.4. Strengths and Limitations

This study has several strengths. It is to our knowledge, the first evaluation of a digitally integrated, automated SMS and voice reminder system for childhood vaccination in Togo, and one of very few studies of this kind conducted in Francophone West Africa. Both service-side (health worker perceptions and operational data) and demand-side (parental perceptions) dimensions were assessed simultaneously, providing a comprehensive feasibility profile of the intervention. However, our study has some limitations. The absence of a control group makes it difficult to attribute the observed changes with certainty to the intervention rather than to other confounding factors (parallel awareness campaigns, general system improvements). The Penta 3 coverage data are derived from the routine administrative health data information (DHIS2) records, the quality and completeness of which may vary from one health facility to another. Furthermore, this is a study conducted exclusively in an urban setting (Grand Lomé), which limits the generalizability of the results to rural areas of Togo.

## 5. Conclusions

This study demonstrates the operational feasibility of deploying a digital reminder system for childhood vaccinations in Togo. The findings support further efforts to optimize the system prior to any consideration of scale-up. In addition, the low marginal cost of SMS and automated calls, combined with the use of existing health system infrastructure, suggests that this intervention could represent a cost-effective complement to traditional outreach strategies.

## Author Contributions

Conceptualization, SB and KMA; methodology, KHG, KMA and ASD; software, KHG; validation, SB, XRS and ECD; formal analysis, KHG; resources, SB, XRS and ECD; data curation, KHG and ASD; writing—original draft preparation, KHG; writing—review and editing, KHG, ASD, SB, XRS and ECD; supervision, SB; project administration, KMA; funding acquisition, SB, XRS and ECD. All authors have read and agreed to the published version of the manuscript.”

## Institutional Review Board Statement

The study was conducted in accordance with the Declaration of Helsinki, and approved by the Institutional Review Board of the School of Health Sciences at the University of Lomé in Togo (N° 409/2026/CE-FSS/19/01).

## Informed Consent Statement

Informed consent was obtained from all subjects involved in the study.

## Data Availability Statement

Data presented in this study are available on request from the corresponding author.

## Acknowledgments

The authors would like to thank all participants in this study.

